# A County-level Analysis of the Association Between Medicaid Expansion and Medicare Spending

**DOI:** 10.1101/2022.08.04.22278455

**Authors:** Walid G. El-Nahal, Mattew D. Eisenberg

## Abstract

**Background:** If access to Medicaid improves health outcomes, it may also result in lower long-term spending, however the association between Medicaid expansion and Medicare spending is unknown. In this analysis we sought to investigate the association between Medicaid expansion and per capita Medicare spending at the county level.

**Methods:** This is an observational analysis of all U.S. counties in the ten years from 2010 to 2019. We used a difference-in-difference event study to investigate the difference in per capita Medicare spending between counties in states that expanded Medicaid and counties in states that did not expand Medicaid. The exposure was treatment year, which characterized whether a county was in an expansion state and when expansion occurred. In non-expansion counties, treatment year was assigned 0 for all observations. In expansion counties, treatment year ranged from -3 to +6, with treatment year 1 corresponding to the first full year of expansion. The primary outcome was fee-for-service Medicare spending per capita in each county. A secondary analysis investigated subcategories of per capita spending including inpatient, outpatient, skilled nursing care, inpatient rehabilitation, home health, and hospice care.

**Results:** We analyzed 1,648 expansion and 1,494 non-expansion counties, with ten observations per county, one for each year between 2010 and 2019. In the adjusted event study analysis, the difference between expansion and non-expansion counties in expansion year 5 compared to pre-expansion was -200 [95% Confidence Interval (CI): -406, 6] dollars. In the subcategory analysis, the difference in inpatient care, skilled nursing care, outpatient care, and home health spending were -46 [95% CI: -103, 12], -92 [95% CI: -194, 11], 57 [95% CI: -67, 181], and 55 [95% CI: -17, 126] dollars per capita respectively.

**Conclusions:** Medicaid expansion is not consistently significantly associated with lower Medicare spending compared to pre-expansion. However, observed trends towards lower spending and cost-shifting from inpatient to outpatient settings in expansion counties warrant additional investigation.

## Introduction

The expansion of Medicaid to all individuals with income below 138% of the Federal Poverty Level (FPL)^1^ through the Patient Protection and Affordable Care Act (ACA) has significantly contributed to lower uninsurance rates in the United States, from 48.3 million in 2010 to 26.4 million in 2022.^2,3^ Following the 2012 Supreme Court ruling in *NFIB v. Sebelius*, the decision to expand Medicaid was left to individual states.^4^ As of this writing, 38 states and the District of Columbia have opted to expand.^5^ The uninsurance rate is nearly twice as high (17.1% vs. 9.1%) in non-expansion states.^6^ Approximately 2.2 million additional adults would be eligible for Medicaid if the remaining states were to expand, more than 75% of whom are in just four states: North Carolina, Georgia, Florida, and Texas.^7^

A significant body of literature exists around investigating the association between Medicaid expansion and patient health.^1,8–16^ Health insurance is a critical component of access to care, with studies demonstrating that insured patients are more likely to seek care.^12,13,17,18^ While insurance has been shown to reduce the financial burden on patients seeking care,^9^ evidence around its effect on health outcomes has been outcome-dependent.^8^ Several studies have shown Medicaid to be associated with improved outcomes including earlier diabetes detection,^13^ better diabetes control,^15^ lower rates of diabetic complications,^16^ less frequent depression,^13^ earlier cancer diagnoses,^19–21^ and improved renal^22,23^ and cardiovascular outcomes,^11^ with some even showing improved mortality.^10^ However, these health improvements have not been universal, with studies of different outcomes showing no significant improvement with Medicaid expansion.^13,24^

If Medicaid does indeed improve some health outcomes, we hypothesize that Medicaid beneficiaries may utilize less healthcare resources in later life once they have aged into Medicare. Whether access to Medicaid does ultimately reduce Medicare spending however is unknown. In this analysis we aim to investigate this, by studying the association between Medicaid expansion and county-level per capita Medicare spending.

## Methods

### Data Sources

We used data on Medicare spending for each county through the Medicare Geographic Variation file from the Centers for Medicare & Medicaid Services (CMS). The contents and methodology for this file are publicly described.^25^ It includes data describing Medicare fee-for-service per capita spending, average beneficiary age, race, ethnicity, gender, and Medicaid eligibility distributions of fee-for-service beneficiaries in each county. Data on healthcare resources including number of hospital beds, skilled nursing beds, and physicians per capita were obtained from the Area Health Resources File compiled by the Health Research & Services Administration.^26^

### Study Sample

Our sample consisted of county-year level-data of 3,142 counties in the 50 states. Annual per capita Medicare fee-for-service spending data from 2010 to 2019 was calculated, resulting in 10 observations per county. In Louisiana and Alaska, where there are no counties, parishes and boroughs respectively were used as county equivalents. Independent cities (e.g. Baltimore City) were also used as county equivalents. The District of Columbia and United States territories were excluded from the analysis.

### Outcome of interest

The outcome of interest was the standardized Medicare fee-for-service spending per enrollee in each county in each year. Standardized spending accounts for geographic variations in costs and is included in the Medicare Geographic Variation file. The methodology for standardization is described elsewhere.^25^ Per capita spending was used to account for differences in population.

### Exposure

The exposure of interest was Medicaid expansion status. The expansion status was manually assigned to counties based on the state they are in and the year the state expanded Medicaid (if ever). Counties were assigned their first expansion year based on the first full year of Medicaid expansion, described in Table 1. The implementation of expansion status in the event study is described in the Statistical Analysis.

**Table 1.**
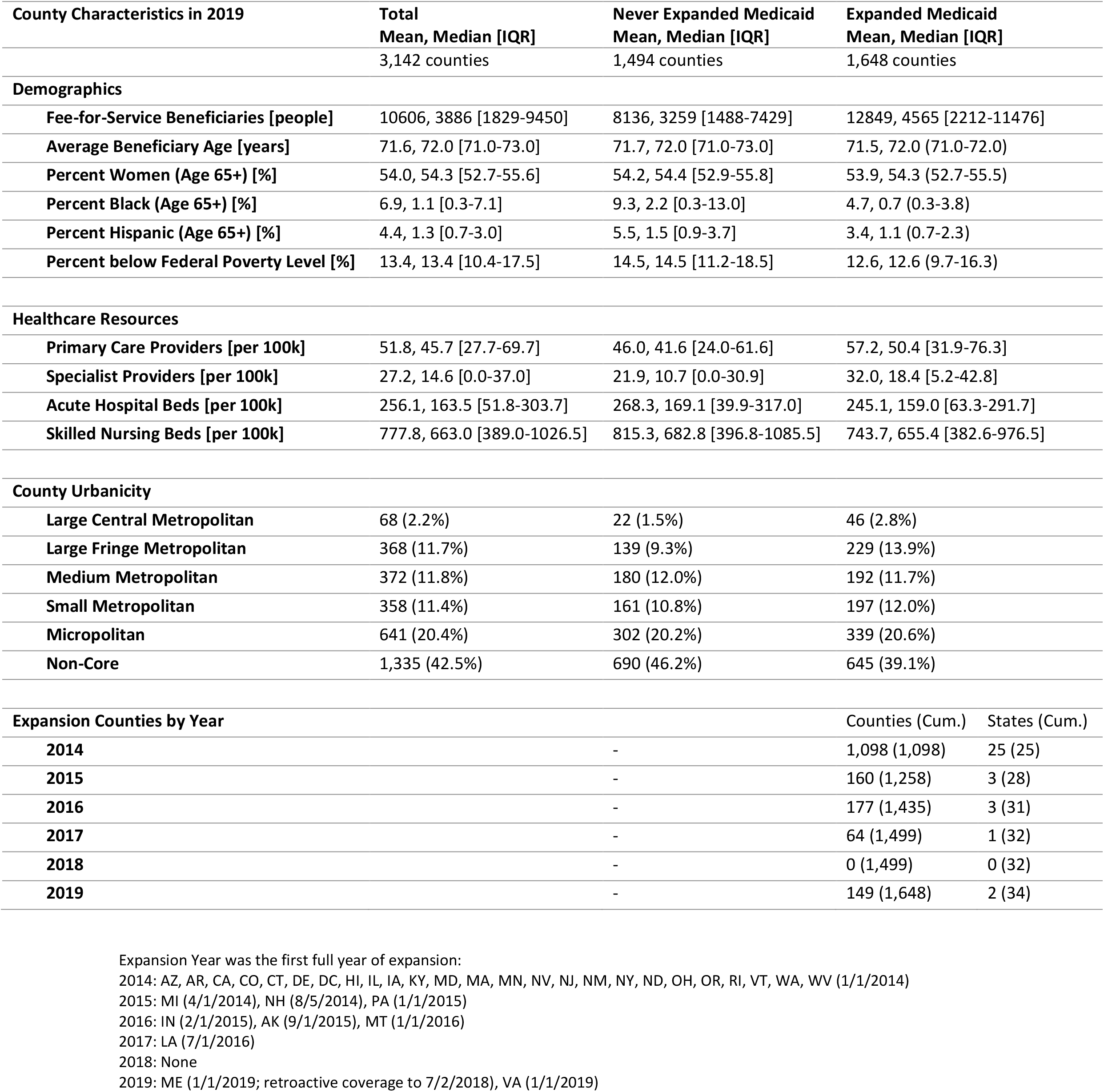
County Characteristics in 2019 by County Expansion Status.

### Covariates

We report difference-in-difference estimators from unadjusted and adjusted models. The covariates of interest in adjusted models were factors that may impact the health of beneficiaries. We used demographic covariates including the average age of Medicare fee-for-service beneficiaries and the proportion of beneficiaries of a given race, ethnicity, and gender. We included the proportion of individuals living in poverty in each county, as this correlates with the proportion of Medicaid eligible beneficiaries. To account for access to healthcare resources, we included the number of per capita acute care hospital beds, skilled nursing beds, primary care physicians, and specialist physicians. All models included variables for state and year fixed effects and were clustered by state.

### Statistical Analysis

We used difference-in-difference^11,25–30^ methods to compare how our outcomes changed over time between populations that did and did not receive an intervention. We used this method to determine whether a change in our outcomes was truly associated with the intervention or merely a product of an ongoing trend.^27^ Similar methods have been extensively used to study the effects of the ACA Medicaid expansion on other outcomes. ^11,15,28–30^ Our analysis used multivariate linear regression accounting for state and calendar year fixed effects. We used an event study or dynamic difference-in-difference design.^31^

An event study analysis, the implementation of which has been described elsewhere,^32^ allows for different intervention times in different areas, which was appropriate given the variable timing of Medicaid expansion across states. For this model, we created a treatment year variable which ranges from -8 to +6 based on when a state expanded. For example, Baltimore County, Maryland was in treatment year -3 in 2010, treatment year 0 in 2013, and treatment year +6 in 2019, because Maryland expanded Medicaid in 2014. Non-expansion states had 0 as their treatment year for all observations. Using this treatment year variable, ten dummy variables were created, designating each possible treatment year. The model included nine of these ten variables as separate terms, resulting in nine difference-in-difference estimators, one for each treatment year, allowing for examination of the magnitude of the effect over time. The dummy variable corresponding to treatment year -1 was omitted from the model to capture the baseline difference between control and treated counties prior to expansion of Medicaid.^32^ In a traditional event study analysis, all observations for all treatment years are included, which was the case for our primary analysis. We also conducted a sensitivity analysis, balancing observations around the treatment year, which restricts to treatment years closer to zero (5 lag and 4 lead years) such that there is a comparable number of pre- and post-treatment observations.

### Secondary Analysis

In the primary analysis, we utilize total Medicare spending. We repeat these analyses using 6 difference categories of spending to determine what type of spending contributed most to the changes. These categories of spending were: inpatient spending, outpatient spending, skilled nursing facility spending, inpatient rehabilitation spending, home health spending, and hospice spending, each per capita.

This work was reviewed by the Johns Hopkins Bloomberg School of Public Health Institutional Review Board (FWA 00000287) and deemed to not qualify as human subjects research and not require review board oversight (IRB00020671). All analyses were conducted in Stata 16.1^33^ using the *eventDD* command.^32^

## Results

The analysis included 1,648expansion and 1,494 non-expansion counties, with ten observations per county, one for each year between 2010 and 2019. County characteristics (Table 1) among Medicare beneficiaries in each county included: mean of 10,607 fee-for-service beneficiaries, mean age 71.6 years, mean 6.9% percent Black population, mean 4.4% percent Hispanic population, mean 54% percent female population. The mean number of primary care providers, specialists, acute hospital beds, and skilled nursing beds were 51.8, 27.2, 256.1, and 777.8 per 100,000 county residents respectively.

In the adjusted event study analysis (Table 2), the difference between expansion and non-expansion counties in treatment year 5 compared to pre-treatment was -200 [CI: -406, 6] dollars. In the secondary analysis, this difference-in-difference coefficient for treatment year 5 in inpatient care, skilled nursing care, outpatient care, and home health were -46 [CI: -103, 12], -92 [CI: -194, 11], 57 [CI: -67, 181], and 55 [CI: -17, 126] dollars respectively.

**Table 2.** Difference-in-Difference in Medicare Fee-for-Service Spending (Dollars Per Enrollee) in Medicaid Expansion Counties compared to those without Medicaid Expansion.

| *Treatment Year* | **Unadjusted Primary** | **Adjusted Primary** | **Adjusted Inpatient** | **Adjusted Outpatient** | **Adjusted SNF** | **Adjusted Rehab** | **Adjusted Home Hlth** | **Adjusted Hospice** |
| --- | --- | --- | --- | --- | --- | --- | --- | --- |
| *Observations<sup>a</sup>* | 31,395 | 31,358 | 31,315 | 31,355 | 30,745 | 23,207 | 30,172 | 29,322 |
| -4 | 49 | 69 | -11 | -59 | 72 | 6 | -3 | 3 |
|  | (-145, 244) | (-92, 231) | (-45, 24) | (-172, 53) | (-0, 144) | (-19, 31) | (-80, 74) | (-19, 25) |
| -3 | -20 | -7 | -10 | -16 | 37** | 6 | -32 | 3 |
|  | (-135, 94) | (-115, 101) | (-48, 28) | (-79, 47) | (5, 70) | (-5, 17) | (-69, 4) | (-9, 14) |
| -2 | -13 | -9 | -12 | -16 | 29** | -0 | -22** | 1 |
|  | (-77, 51) | (-70, 53) | (-31, 7) | (-55, 22) | (9, 50) | (-8, 7) | (-37, -7) | (-5, 7) |
| 0 | -15 | -18 | -19 | -10 | 1 | -9** | 8 | 2 |
|  | (-61, 31) | (-67, 31) | (-39, 2) | (-32, 11) | (-22, 23) | (-18, -1) | (-6, 22) | (-6, 11) |
| +1 | -75** | -80** | -21** | -4 | -15 | -17** | 16 | 1 |
|  | (-143, -6) | (-152, -8) | (-42, -1) | (-55, 46) | (-52, 21) | (-31, -3) | (-8, 40) | (-14, 15) |
| +2 | -122** | -122** | -34** | 21 | -36 | -22 | 14 | -9 |
|  | (-223, -21) | (-225, -18) | (-63, -5) | (-40, 83) | (-93, 21) | (-44, 0) | (-31, 59) | (-26, 8) |
| +3 | -133 | -132 | -33** | 44 | -54 | -30** | 16 | -9 |
|  | (-271, 5) | (-268, 5) | (-67, -0) | (-38, 126) | (-128, 20) | (-58, -2) | (-39, 71) | (-28, 10) |
| +4 | -123 | -117 | -23 | 69 | -69 | -37** | 46 | -13 |
|  | (-290, 45) | (-285, 52) | (-66, 21) | (-30, 167) | (-164, 25) | (-73, -1) | (-14, 105) | (-35, 8) |
| +5 | -214** | -200 | -46 | 57 | -92 | -45 | 55 | -13 |
|  | (-416, -12) | (-406, 6) | (-103, 12) | (-67, 181) | (-194, 11) | (-90, 1) | (-17, 126) | (-39, 12) |

\*\*p<0.05. Difference-in-difference Coefficient (Confidence Interval).
Units are dollars per capita.
<sup>a</sup>Treatment year -1 was omitted from the model, to capture the baseline difference between control and treated counties prior to expansion of Medicaid
<sup>b</sup>There are <31,420 observations (10 observations of 3,142 counties) due to missingness of data points.

## Discussion

The main finding of this analysis is that Medicaid expansion in the first 5 years was not consistently significantly associated with lower Medicare spending compared to pre-expansion, however we did observe a demonstrable trend towards lower spending, with two post-treatment years reaching significance. Additionally, the secondary analysis demonstrated trends of decreased Medicare spending on inpatient acute care, skilled nursing facilities and rehabilitation hospitals, and trends of increased Medicare spending on outpatient care and home health with Medicaid expansion. The observation of trends without consistent statistical significance suggests that the analysis of 3,143 counties may have been underpowered to detect the effect under study, and that future work to further explore this association is warranted.

Our initial hypothesis was that access to Medicaid may result in reduced Medicare spending per capita in the long run, following the hypothesis that Medicaid expansion would offer eligible patients access to consistent long-term care, allowing for better management of chronic conditions. This is reflected in existing literature demonstrating that Medicaid expansion was associated with improved outcomes of several chronic disease including diabetes,^13,15,16^ kidney disease,^22,23^ cardiovascular disease,^11^ mental health disorders,^13^ and cancer.^19–21^ In turn, our expectation was that this improved control of chronic conditions would result in less frequent spending on catastrophic care, such as hospitalizations for life-threatening and costly complications of diabetes, hypertension, heart disease, etc. While our findings were not statistically different from zero, the observed trends are consistent with the hypothesis. Furthermore, a sensitivity analysis of balanced treatment years, years with comparable numbers of pre-expansion and post-expansion observations, demonstrated a significant decrease in per capita Medicare spending associated with Medicaid expansion.

The secondary analysis also demonstrated trends of decreased inpatient spending on acute care, inpatient rehabilitation, and skilled nursing. In contrast, we observed increased spending on outpatient care and home health. This shift in cost from inpatient settings to outpatient settings would also be consistent with the hypothesis that Medicaid-eligible patients benefit from access to outpatient care, which may continue into their Medicare years and mitigate the risk and cost of hospitalizations for acute events. Interestingly, prior literature has suggested that Medicaid access is associated with an increase in utilization of long-term care,^34^ including home health which is consistent with our findings, but also skilled nursing which was inconsistent with our findings. This could suggest that reduced Medicare spending on skilled nursing is merely an artifact of shifting the burden to Medicaid, which is an important source of skilled nursing reimbursement. Future work is warranted to investigate how Medicaid expansion impacts the distribution of expenses, to better characterize its economic impact on our healthcare system.

Several limitations of this analysis are worth noting. This is a county-level analysis of per capita spending at a population level. While we adjusted for available covariates including population demographics and social determinants, these covariates are unlikely to account for all factors that influence Medicare spending, such as individual comorbidity. County-level analyses are also limited to 3,142 observations, which restricts the power of the analysis to detect smaller differences in spending, and may explain the observation of trends that in some cases did not reach statistical significance. Future individual-level analyses are needed to build on this work. Additionally, costs do vary across the country. We used standardized costs to mitigate this effect, but it likely has some influence on the outcome. Future work is also needed to assess the trends studied herein over longer timescales. If Medicaid does improve health in a way that ultimately results in lower long-term healthcare costs, such a phenomenon would likely be better observed over a longer time scale than the 5-year post-expansion period studied in this analysis. Finally, dynamic difference-in-difference or event study analyses have known limitations, including reliance on pre-treatment parallel trends and assumptions about homogeneity of the treatment effects, which limit the strength of inference.^35,36^ Regardless of these limitations however, this analysis does demonstrate that further investigations into the impact of Medicaid expansion on Medicare spending are warranted to determine if Medicaid may meaningfully curb long-term care costs.

## Conclusion

In summary, Medicaid expansion is not significantly associated with reduced Medicare spending, but there is a demonstrable trend in that direction, which appears to be driven by trends towards shifting expenses from inpatient care to outpatient care. While this work does not definitively demonstrate an association, it holds value as a means of generating hypotheses around the link between access to insurance and long-term healthcare costs. Future work is needed to re-evaluate this trend as more patients with access to Medicaid expansion age into Medicare, and individual-level investigations would be of particular value. If future analyses bear out the trends observed here, they may have significant implications on the importance of expanding insurance access on health and healthcare spending.

**Figure 1.**
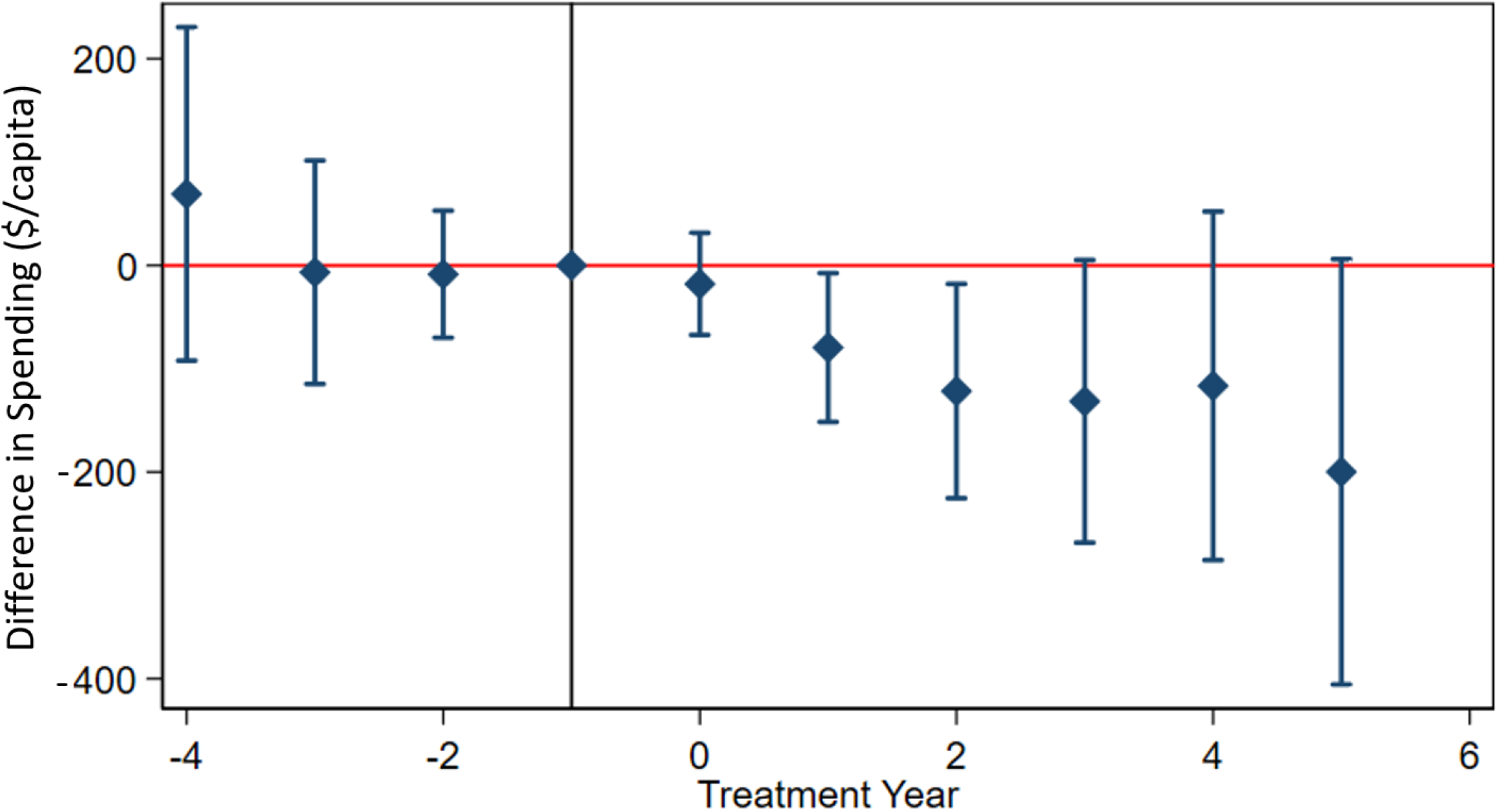
Difference in Medicare Fee-for-Service Spending Per Enrollee Between Expansion and Non-Expansion Counties by Treatment Year. Difference-in-difference (event study) analysis of Medicare fee-for-service spending per enrollee in expansion vs non-expansion counties by treatment year. Includes year and state fixed effects and is adjusted for average age and proportions of race, ethnicity, and gender of beneficiaries, county poverty rates, and per capita hospital beds, skilled nursing beds, primary care physicians, and specialists.

**Figure 2.**
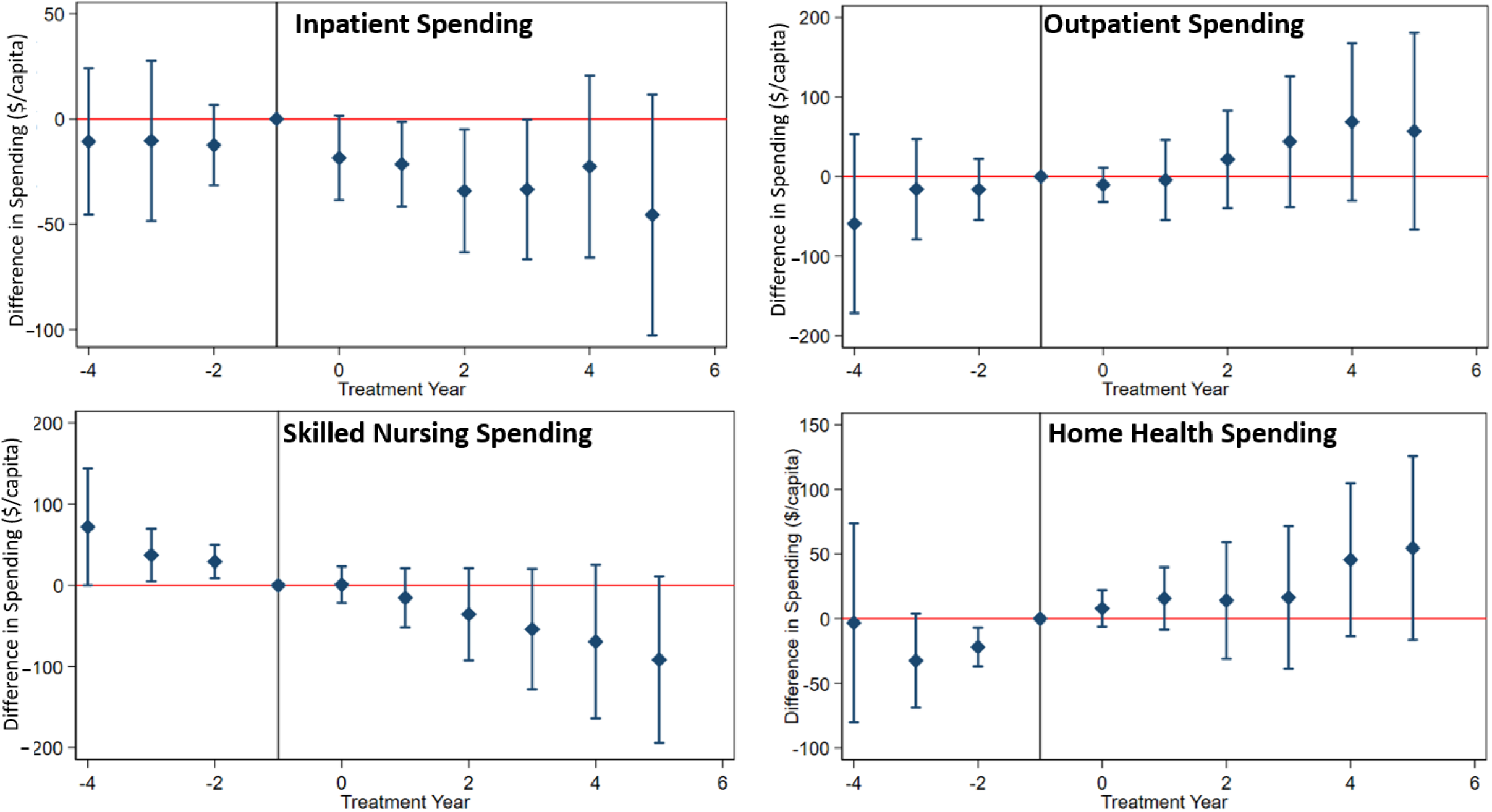
Difference in Medicare Fee-for-Service Spending Per Enrollee Between Expansion and Non-Expansion Counties by Treatment Year and Spending Type. Difference-in-difference (event study) analysis of Medicare fee-for-service spending per enrollee by spending category in expansion vs non-expansion counties across treatment years. Includes year and state fixed effects and is adjusted for average age and proportions of race, ethnicity, and gender of beneficiaries, county poverty rates, and per capita hospital beds, skilled nursing beds, primary care physicians, and specialists.

## Data Availability

All data produced in the present study are available upon reasonable request to the authors.

https://data.cms.gov/summary-statistics-on-use-and-payments/medicare-geographic-comparisons/medicare-geographic-variation-by-national-state-county

https://data.hrsa.gov/topics/health-workforce/ahrf

## SUPPLEMENTAL FIGURES

**Supplemental Figure 1.**
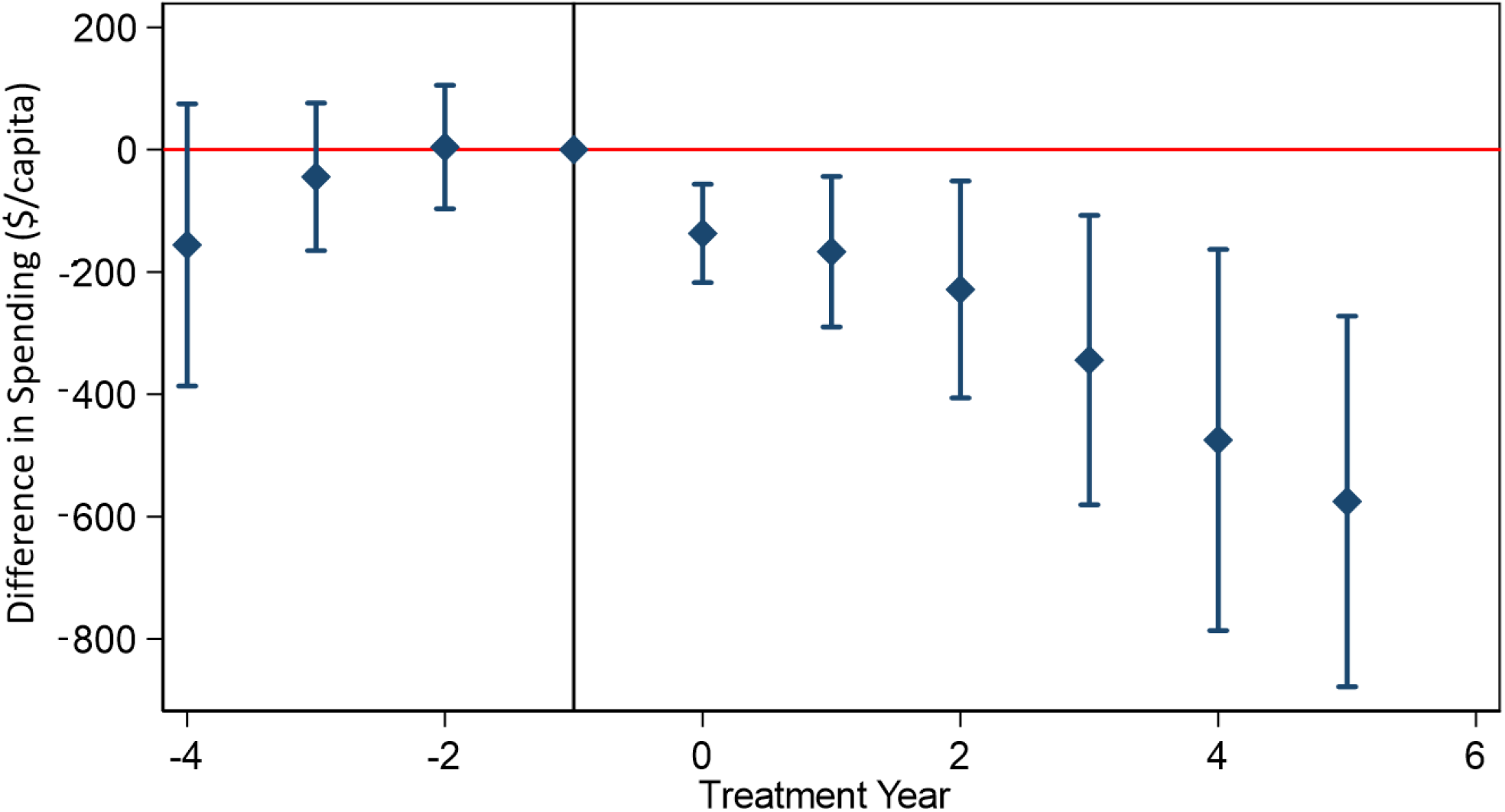
Difference in Medicare Fee-for-Service Spending Per Enrollee Between Expansion and Non-Expansion Counties across Balanced Treatment Years. Balanced difference-in-difference (event study) analysis of Medicare fee-for-service spending per enrollee by spending category in expansion vs non-expansion counties across treatment years. Balanced analysis includes observations from 5 lag and 4 lead years. Model includes year and state fixed effects and is adjusted for average age and proportions of race, ethnicity, and gender of beneficiaries, county poverty rates, and per capita hospital beds, skilled nursing beds, primary care physicians, and specialists.

## References

1. Buchmueller TC, Levy HG. The ACA’s impact on racial and ethnic disparities in health insurance coverage and access to care. Health Affairs. 2020;39(3):395–402. doi:10.1377/hlthaff.2019.01394

2. Blumenthal D, Collins SR, Fowler EJ. The Affordable Care Act at 10 Years — Its Coverage and Access Provisions. New England Journal of Medicine. 2020;382(10):963–969. doi:10.1056/NEJMHPR1916091/SUPPL_FILE/NEJMHPR1916091_DISCLOSURES.PDF

3. Lee A, Ruhter J, Peters C, de Lew N, Sommers BD. National Uninsured Rate Reaches All-Time Low in Early 2022.; 2022. Accessed August 2, 2022. https://aspe.hhs.gov/sites/default/files/documents/15c1f9899b3f203887deba90e3005f5a/Uninsured-Q1-2022-Data-Point-HP-2022-23-08.pdf

4. Oberlander J. The Ten Years’ War: Politics, Partisanship, And The ACA. https://doi-org.proxy1.library.jhu.edu/101377/hlthaff201901444. 2020;39(3):471–478. doi:10.1377/HLTHAFF.2019.01444

5. The Kaiser Family Foundation. Status of State Medicaid Expansion Decisions: Interactive Map. Accessed May 5, 2021. https://www.kff.org/medicaid/issue-brief/status-of-state-medicaid-expansion-decisions-interactive-map/

6. Finegold K, Conmy A, Chu RC, Bosworth A, Sommers BD. Trends in the U.S. Uninsured Population, 2010-2020.; 2010.

7. Levitt L. The Inequity of the Medicaid Coverage Gap and Why It Is Hard to Fix It. JAMA Health Forum. 2021;2(10):e213905–e213905. doi:10.1001/JAMAHEALTHFORUM.2021.3905

8. Soni A, Wherry LR, Simon KI. How have ACA insurance expansions affected health outcomes? Findings from the literature. Health Affairs. 2020;39(3):371–378. doi:10.1377/hlthaff.2019.01436

9. Glied SA, Collins SR, Lin S. Did the ACA Lower Americans’ Financial Barriers to Health Care? Health Affairs. 2020;39(3):379–386. doi:10.1377/hlthaff.2019.01448

10. Miller S, Johnson N, Wherry L. Medicaid and Mortality: New Evidence from Linked Survey and Administrative Data.; 2019. doi:10.3386/w26081

11. Khatana SAM, Bhatla A, Nathan AS, et al. Association of Medicaid Expansion with Cardiovascular Mortality. JAMA Cardiology. 2019;4(7):671–679. doi:10.1001/jamacardio.2019.1651

12. Miller S, Wherry LR. Health and Access to Care during the First 2 Years of the ACA Medicaid Expansions. New England Journal of Medicine. 2017;376(10):947–956. doi:10.1056/NEJMSA1612890/SUPPL_FILE/NEJMSA1612890_DISCLOSURES.PDF

13. Baicker K, Taubman SL, Allen HL, et al. The Oregon Experiment — Effects of Medicaid on Clinical Outcomes. New England Journal of Medicine. 2013;368(18):1713–1722. doi:10.1056/NEJMSA1212321/SUPPL_FILE/NEJMSA1212321_DISCLOSURES.PDF

14. Allen H, Sommers BD. Medicaid Expansion and Health: Assessing the Evidence After 5 Years. JAMA. 2019;322(13):1253–1254. doi:10.1001/JAMA.2019.12345

15. Cole MB, Kim JH, Levengood TW, Trivedi AN. Association of Medicaid Expansion With 5-Year Changes in Hypertension and Diabetes Outcomes at Federally Qualified Health Centers. JAMA Health Forum. 2021;2(9):e212375–e212375. doi:10.1001/JAMAHEALTHFORUM.2021.2375

16. Tan TW, Calhoun EA, Knapp SM, et al. Rates of Diabetes-Related Major Amputations Among Racial and Ethnic Minority Adults Following Medicaid Expansion Under the Patient Protection and Affordable Care Act. JAMA Network Open. 2022;5(3):e223991–e223991. doi:10.1001/JAMANETWORKOPEN.2022.3991

17. Brook RH, Keeler EB, Lohr KN, et al. The Health Insurance Experiment A Classic RAND Study Speaks to the Current Health Care Reform Debate Key Fi Ndings. RAND Corporation; 2006.

18. Aron-Dine A, Einav L, Finkelstein A. The RAND health insurance experiment, three decades later. Journal of Economic Perspectives. 2013;27(1):197–222. doi:10.1257/jep.27.1.197

19. Han X, Yabroff KR, Ward E, Brawley OW, Jemal A. Comparison of Insurance Status and Diagnosis Stage Among Patients With Newly Diagnosed Cancer Before vs After Implementation of the Patient Protection and Affordable Care Act. JAMA oncology. 2018;4(12):1713–1720. doi:10.1001/JAMAONCOL.2018.3467

20. Lin L, Soni A, Sabik LM, Drake C. Early- and Late-Stage Cancer Diagnosis Under 3 Years of Medicaid Expansion. American journal of preventive medicine. 2021;60(1):104–109. doi:10.1016/J.AMEPRE.2020.06.020

21. Lam MB, Phelan J, Orav EJ, Jha AK, Keating NL. Medicaid Expansion and Mortality Among Patients With Breast, Lung, and Colorectal Cancer. JAMA Network Open. 2020;3(11):e2024366–e2024366. doi:10.1001/JAMANETWORKOPEN.2020.24366

22. Swaminathan S, Sommers BD, Thorsness R, Mehrotra R, Lee Y, Trivedi AN. Association of Medicaid Expansion With 1-Year Mortality Among Patients With End-Stage Renal Disease. JAMA. 2018;320(21):2242–2250. doi:10.1001/JAMA.2018.16504

23. Thorsness R, Swaminathan S, Lee Y, et al. Medicaid Expansion and Incidence of Kidney Failure among Nonelderly Adults. Journal of the American Society of Nephrology. 2021;32(6):1425–1435. doi:10.1681/ASN.2020101511

24. Soni A, Wherry LR, Simon KI. How have ACA insurance expansions affected health outcomes? Findings from the literature. Health Affairs. 2020;39(3):371–378. doi:10.1377/hlthaff.2019.01436

25. Medicare Geographic Variation - by National, State & County - Centers for Medicare & Medicaid Services Data. data.cms.gov.

26. Area Health Resources Files. Accessed March 16, 2022. https://data.hrsa.gov/topics/health-workforce/ahrf

27. Dimick JB, Ryan AM. Methods for Evaluating Changes in Health Care Policy: The Difference-in-difference Approach. JAMA. 2014;312(22):2401–2402. doi:10.1001/JAMA.2014.16153

28. Graves JA, Fry C, McWilliams JM, Hatfield LA. Difference-in-difference for categorical outcomes. Health Services Research. 2022;57(3):681–692. doi:10.1111/1475-6773.13948

29. Courtemanche C, Marton J, Ukert B, Yelowitz A, Zapata D. Effects of the Affordable Care Act on Health Care Access and Self-Assessed Health After 3 Years. Inquiry: A Journal of Medical Care Organization, Provision and Financing. 2018;55. doi:10.1177/0046958018796361

30. Griffith K, Evans L, Bor J. The affordable care act reduced socioeconomic disparities in health care access. Health Affairs. 2017;36(8):1503–1510. doi:10.1377/HLTHAFF.2017.0083/ASSET/IMAGES/LARGE/2017.0083FIGEX4.EPS.GZ.JPEG

31. Marcus M, Sant’Anna PHC. The Role of Parallel Trends in Event Study Settings: An Application to Environmental Economics. Journal of the Association of Environmental and Resource Economists. 2021;8(2):235–275. doi:10.1086/711509/ASSET/IMAGES/LARGE/FG5.JPEG

32. Clarke D, Tapia-Schythe K. Implementing the panel event study: https://doi.org/101177/1536867X211063144. 2022;21(4):p853-884. doi:10.1177/1536867X211063144

33. StataCorp. Stata Statistical Software. Release 16.

34. Van Houtven CH, McGarry BE, Jutkowitz E, Grabowski DC. Association of Medicaid Expansion Under the Patient Protection and Affordable Care Act With Use of Long-term Care. JAMA Network Open. 2020;3(10):e2018728–e2018728. doi:10.1001/JAMANETWORKOPEN.2020.18728

35. Roth J. Pre-test with Caution: Event-study Estimates After Testing for Parallel Trends.

36. Sun L, Abraham S. Estimating dynamic treatment effects in event studies with heterogeneous treatment effects. Journal of Econometrics. 2021;225(2):175–199. doi:10.1016/J.JECONOM.2020.09.006

